# The false positive paradox: Examining real-world clinical predictive performance of FDA-authorized AI devices for radiology using clinical prevalence

**DOI:** 10.64898/2026.03.25.26349197

**Authors:** Erin Sparnon, Kelvin Stevens, Elizabeth C. Song, Robert J. Harris, Benjamin W. Strong, Michael A. Bruno, Grayson L. Baird

**Affiliations:** SEI (Systems Evolution, Inc.); Faculty of Medicine, University of Freiburg, Freiburg im Breisgau, Germany; Brown Radiology Human Factors Lab, Providence, RI, USA; The Warren Alpert Medical School, Brown University, Providence, RI, USA; vRad® (Virtual Radiologic); Department of Radiology, Penn State Milton S. Hershey Medical Center, Hershey, PA, USA; Department of Radiology, Brown University Health and The Warren Alpert Medical School, Brown University, Providence, RI, USA

## Abstract

The present study evaluates the real-world clinical predictive performance of FDA-authorized artificial intelligence (AI) devices used in radiology, focusing on the false positive paradox (FPP) and its implications for clinical practice. To do this, we analyzed publicly available FDA data on AI radiology devices from 2024 and 2025 from 510(k) summaries, demonstrating how diagnostic accuracy metrics like sensitivity and specificity do not necessarily translate into high positive predictive value (PPV) due to the influence of target disease prevalence. We show the importance of disclosing the false discovery (FDR) and false omission rates (FOR) and argue that this transparency enables clinicians to select AI systems that balance false positive and false negative costs in a clinically, ethically, and financially appropriate manner. Finally, we provide recommendations for what data should be provided to best serve practices and radiologists.

## Introduction

Artificial intelligence (AI) detection systems, such as radiological computer-assisted triage and notification software, in radiology are generally evaluated based on their diagnostic accuracy, namely sensitivity and specificity. These metrics are commonly disclosed in US Food and Drug Administration (FDA) submissions^1^. Although the FDA does not require specific metrics and thresholds of those metrics, vendors often point to their disclosed sensitivity and specificity as an important selling point and indicator of accuracy. However, even demonstrating high diagnostic accuracy, however defined, does not necessarily guarantee correspondingly high predictive performance, such as positive predictive value (PPV). For example, even after achieving high sensitivity and specificity, an AI system can generate far more false positives than true positives. How can this be?

The reason is that predictive performance, PPV, and negative predictive value (NPV) are a function not only of diagnostic accuracy but also of the prevalence of the diagnostic target ^2^. As such, when the prevalence of the diagnostic target is low, even an AI system with high sensitivity and specificity can incorrectly flag many healthy patients (false positives) along with the few sick patients (true positives). These false alarms are measured as the false discovery rate (FDR, 1-PPV). And vice versa, if the prevalence of the diagnostic target were high (although perhaps rare in practice), even an AI system with high sensitivity and specificity can incorrectly fail to flag many sick patients (false negatives) along with the few healthy patients. These misses are measured as the false omission rate (FOR, 1-NPV). That false positives become more probable than true positives as the prevalence of the target decreases was perhaps first formally documented as the “false-positive paradox” by National Aeronautics and Space Administration (NASA)^3^ and, more broadly, as the “accuracy paradox” ^4^.

The false-positive paradox (FPP) has several potential downstream effects worth considering. The first is that clinicians may not anticipate that FDA-authorized devices demonstrating sufficient accuracy will produce many false positives. This is an example of base rate neglect, a cognitive bias in which individuals ignore statistical information in favor of specific information^5,6^. Here, the target’s prevalence (base rate) is ignored in favor of the AI system’s high sensitivity and specificity (or assumed high accuracy due to FDA authorization). By not understanding the FPP, clinicians’ expectations may be misaligned, leading them to ultimately distrust an AI system because they misperceive that it produces “too many false positives,” as previously documented^7^.

What is more, even if clinicians understand the FPP, they may also regard an abundance of false positives from AI, documented in a patient’s files, as a legal liability—what happens when clinicians (eventually) miss something that AI had “caught”^8^? Experimental jury and patient research only validate these concerns—an AI result detecting abnormality will likely increase the clinician’s liability^9,10^. Thus, clinicians will be incentivized to agree with an AI’s flag for abnormality. Specifically, the cost of disagreeing with AI—legally, financially, and reputationally— could vastly outweigh the cost of agreeing with it. For example, the “better safe than sorry” mentality can lead to ordering follow-up imaging even if it is most often unnecessary, perhaps the hallmark of defensive medicine. Here, for the clinician, the cost of a single false negative (missed pathology) outweighs the cost of many false positives (unnecessary imaging), a phenomenon known as the adaptive bias^8,11^.

In addition, experimental research has demonstrated that false-positive AI outputs can increase a clinician’s likelihood of also calling a case positive, even for cases they correctly ruled out without AI, likely due to anchoring and/or automation bias^8^. Thus, an AI’s high rate of false positives, driven by prevalence rather than accuracy, could subsequently fuel a clinician’s false-positive rate, driven by cognitive biases. Of course, the downstream effects of these false positives can manifest as healthy patients subjected to unnecessary follow-up imaging, invasive biopsies, and anxiety, all while consuming resources that would be better used on sick patients^7,10^.

Finally, the aforementioned concerns may be exacerbated, as vendors sometimes report artificially inflated PPV and deflated NPV using a enriched datasets for testing without adjusting them to real-world prevalence^7^. Theoretically, the PPV and NPV estimates should provide clinicians with the correct error rates (1-PPV=FDR and 1-NPV=FOR). However, case enrichment is often used to test AI systems to increase the stability of estimates of accuracy, namely, sensitivity. Although case enrichment achieves this aim, the dataset’s prevalence is therefore not representative of the real-world population’s prevalence, which is much lower.Thus, PPVs published in public 510(k) summaries can be much higher than clinical practice, and clinicians’ expectations of the FDR and FOR can therefore again be vastly misaligned with what they will encounter in clinical practice^7^.

Fortunately, clinicians can calculate more realistic PPV (and FDR) and NPV (and FOR) values even when using sensitivity and specificity data provided in public 510(k) summaries.Specifically, local practices can estimate their own PPV (and FDR) and NPV (and FOR) for a given AI system using their own prevalence estimates or, if unavailable, prevalence rates from the published literature. The purpose of the present study is to demonstrate this.

## Methods

### Data collection

A cohort of recent radiological computer-assisted triage and notification software (i.e., products codes QAS, QBS, QDQ, QFM) from 2024 and 2025 was collected from the FDA’s 510(k) Premarket Notification Database^12^ on October 21, 2025. Because vendor-reported performance data is generally disclosed as part of public FDA decision summaries, we searched the FDA-published list of “<u>Artificial Intelligence-Enabled Medical Devices”</u>^13^ for systems with

1. “Radiology” as a Lead Panel
2. Date of Final Decision in 2024 or 2025
3. Product code: QAS, QBS, QDQ, QFM

This resulted in a cohort of 38 systems, each with a link to its public premarket decision page. Then two reviewers independently (XX,XX) identified and extracted from these public decision summaries, when available, the following performance metrics for each pathology identified in the systems’ Indications For Use:

1. Pathology name (e.g., Aortic Dissection, Intracranial Hemorrhage)
2. Sensitivity
3. Specificity
4. Receiver operating characteristic Area Under the Curve
5. Prevalence of pathology within the validation data
6. Positive Predictive Value
7. Negative Predictive Value

A third reviewer (XX) identified and resolved any inconsistencies between the two reviewers. The final cohort is listed in Table 1 as described below.

**Table 1.**
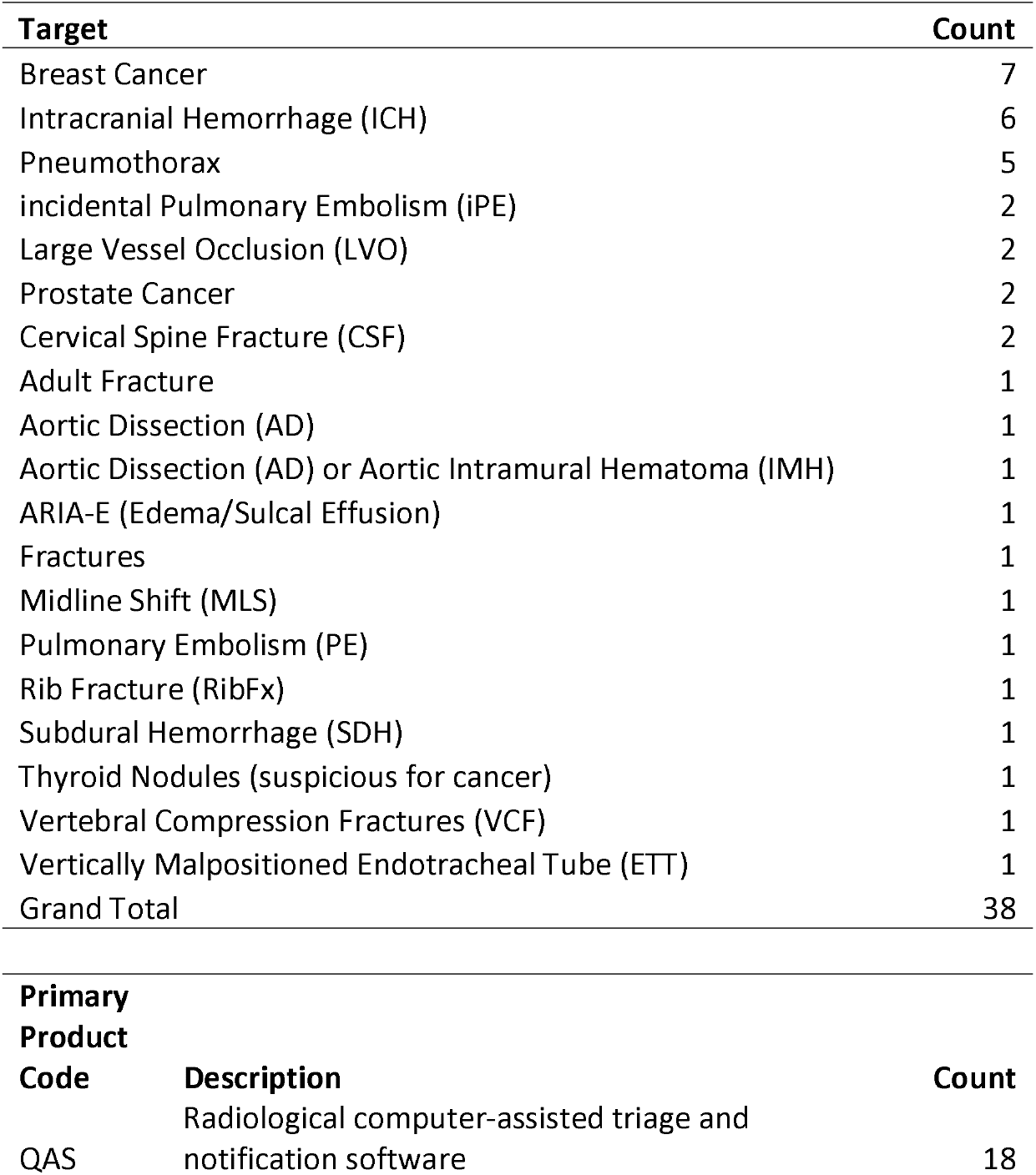
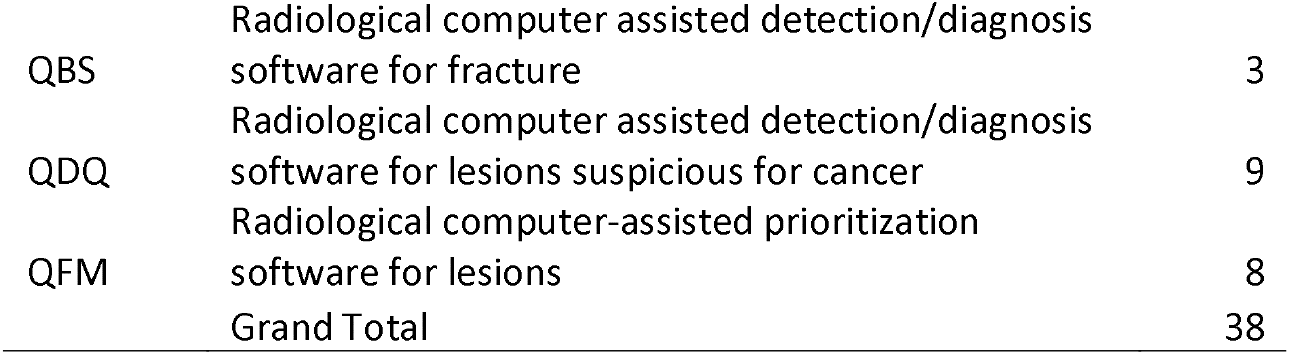
Counts of Targets and Product Types.

### Base rates

Once the devices were identified, the clinical prevalence rates of the diagnostic targets corresponding with these devices were identified either internally by a private practice (XX, details XXX) or from the published literature. Clinical prevalence from the clinical practice was derived as the scan-level incidence: Pulmonary Embolism (2.519%), Aortic Dissection (0.322%), Intracranial Hemorrhage (6.658%), C-Spine Fracture (2.428%), Pneumothorax (1.028%), as seen in Table 2.

**Table 2.** Clinical Prevalence Table.

| Pathology | Total Scans | Scans with Pathology | Scan-Level Incidence (%) |
| --- | --- | --- | --- |
| **Pulmonary Embolism** | 1,074,329 | 27,065 | 2.519 |
| **Aortic Dissection** | 1,686,171 | 5,431 | 0.322 |
| **Intracranial Hemorrhage** | 786,014 | 52,336 | 6.658 |
| **C-Spine Fracture** | 1,066,625 | 25,899 | 2.428 |
| **Pneumothorax** | 1,981,789 | 20,373 | 1.028 |

### Statistical methods

Clinical PPV (and FDR) and NPV (and FOR) were calculated using Bayes’ theorem (see Supplemental Materials, Equations 1-4) using private practice or literature prevalence rates. Overall sensitivity, specificity, and ROC-AUC were estimated using generalized linear mixed modeling (nesting by summary) assuming binomial distribution with SAS Software 9.4 (Cary, NC) using the GLIMMIX procedure. This was done only to describe the overall sensitivity, specificity, and ROC-AUC for the summaries.

## Results

As shown in Table 3, there were 38 devices submitted for one or more target pathologies, for a total of 57 entries. Out of these 57 entries, 91% (52/57) provided sensitivity and specificity, with 5% (3/57) reporting only ROC-AUC (K241747, K242652, K240697) and the remaining two reporting no diagnostic performance metric (K241770, K243688). Of these two, one (K241770) reported the sensitivity, specificity, and ROC-AUC of radiologists using the device, but did not include a standalone analysis of the device. Of the 57, 36 (63%) reported the ROC-AUC along with sensitivity and specificity. The meta-analytic mean of sensitivity, specificity, and ROC-AUC were 92.6 (95% CI [91.2, 93.9]), 90.8 (95% CI [87.5, 93.4]), and 95.2 (95% CI [92.7, 96.9]),respectively.

**Table 3.**
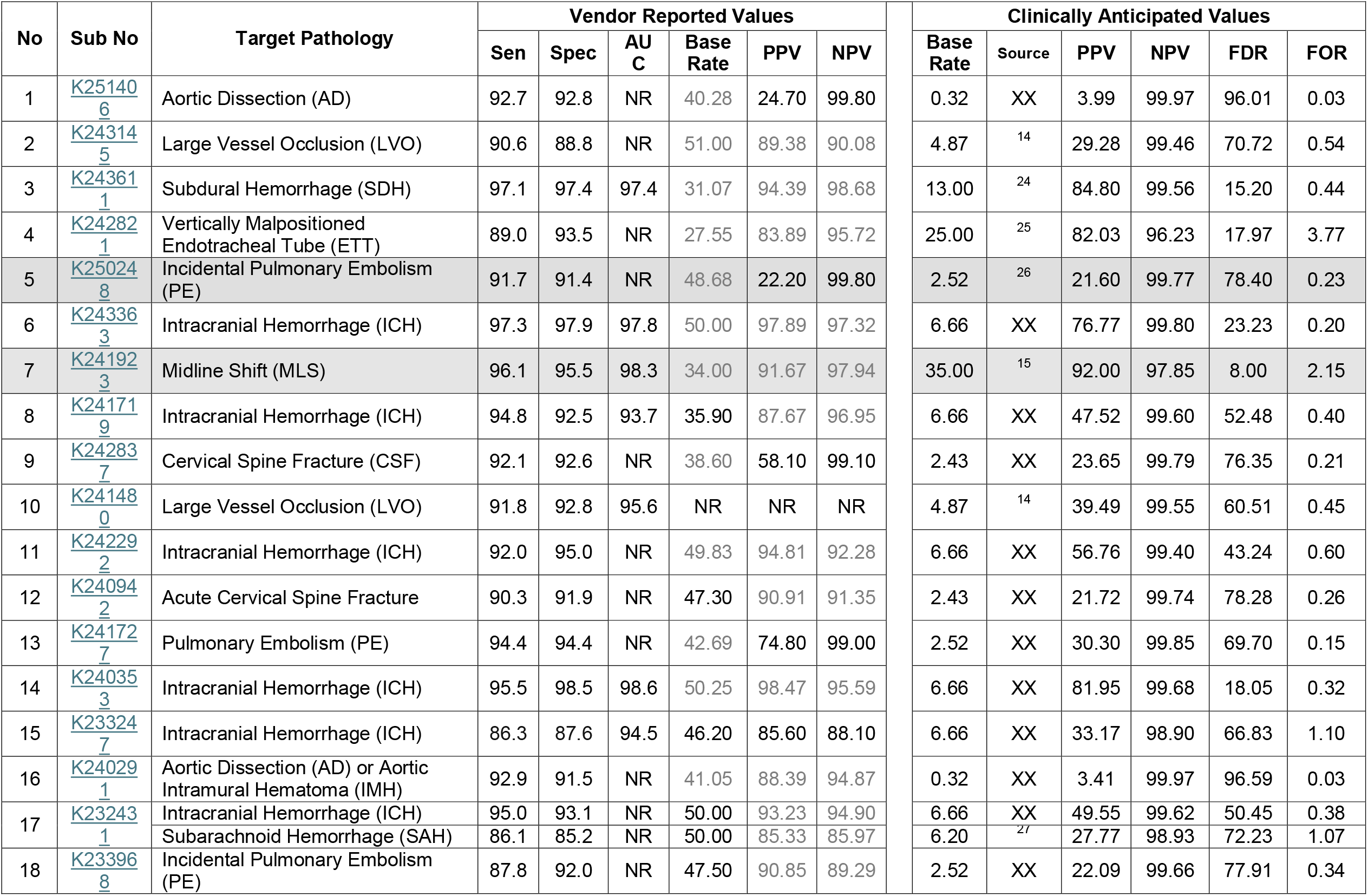
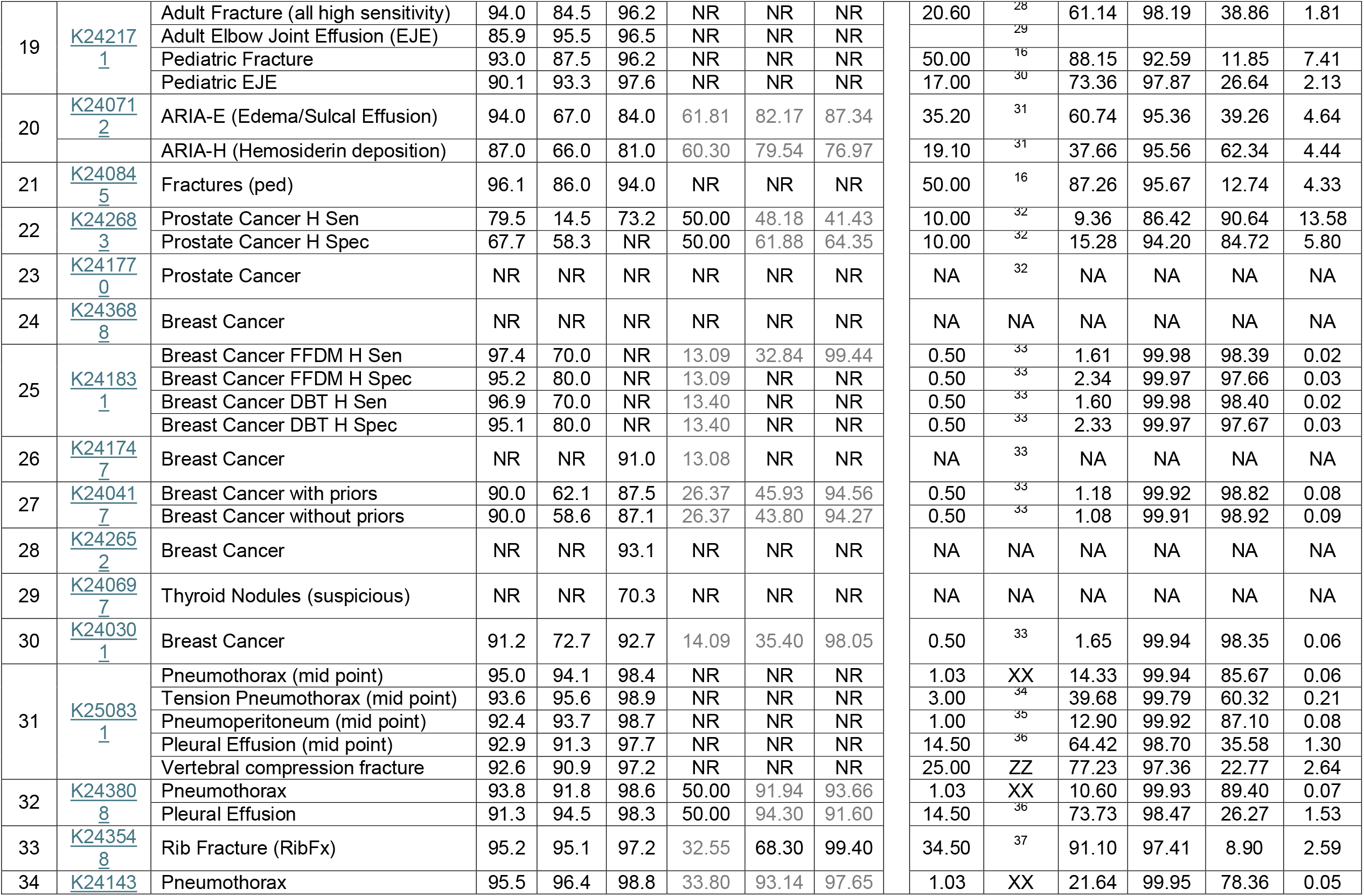
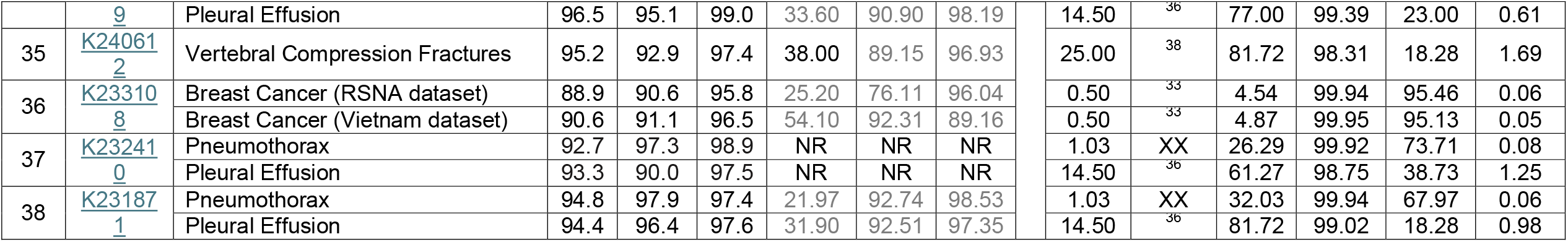
Metrics by Summary number. Note: Sen=sensitivity; Spec=specificity; AUC=Area under the curve; PPV=positive predictive value; NPV=negative predictive value; FDR=false discovery rate; FOR=false omission rate; NR=not reported; Gray text indicates amount was derived from the data available. Source is XX private practice or ZZ derived from citation number. Base rate=prevalence estimate. All numbers are in percentages. Gray highlighted boxes indicate that the PPV and NPV closely reflect clinically anticipated PPV and NPV.

Six public submission summaries (10%) explicitly reported PPV and NPV (black text), and 25 (43%) reported enough data to derive the PPV and NPV (gray text). Moreover, 11 (19%) summaries explicitly reported prevalence (black text), and 29 (51%) implicitly reported it (gray text). Overall means were not calculated for PPV, NPV, or prevalence, as these are specific to pathology and will vary greatly as a function of prevalence. However, as illustrated in Table 3, when PPV and NPV are calculated using 510(k) summary sensitivity and specificity along with prevalence from a clinical practice or the literature, the FDR is often very high, exceeding 50%, especially for low-prevalence pathology (Figure 1).

**Figure 1.**
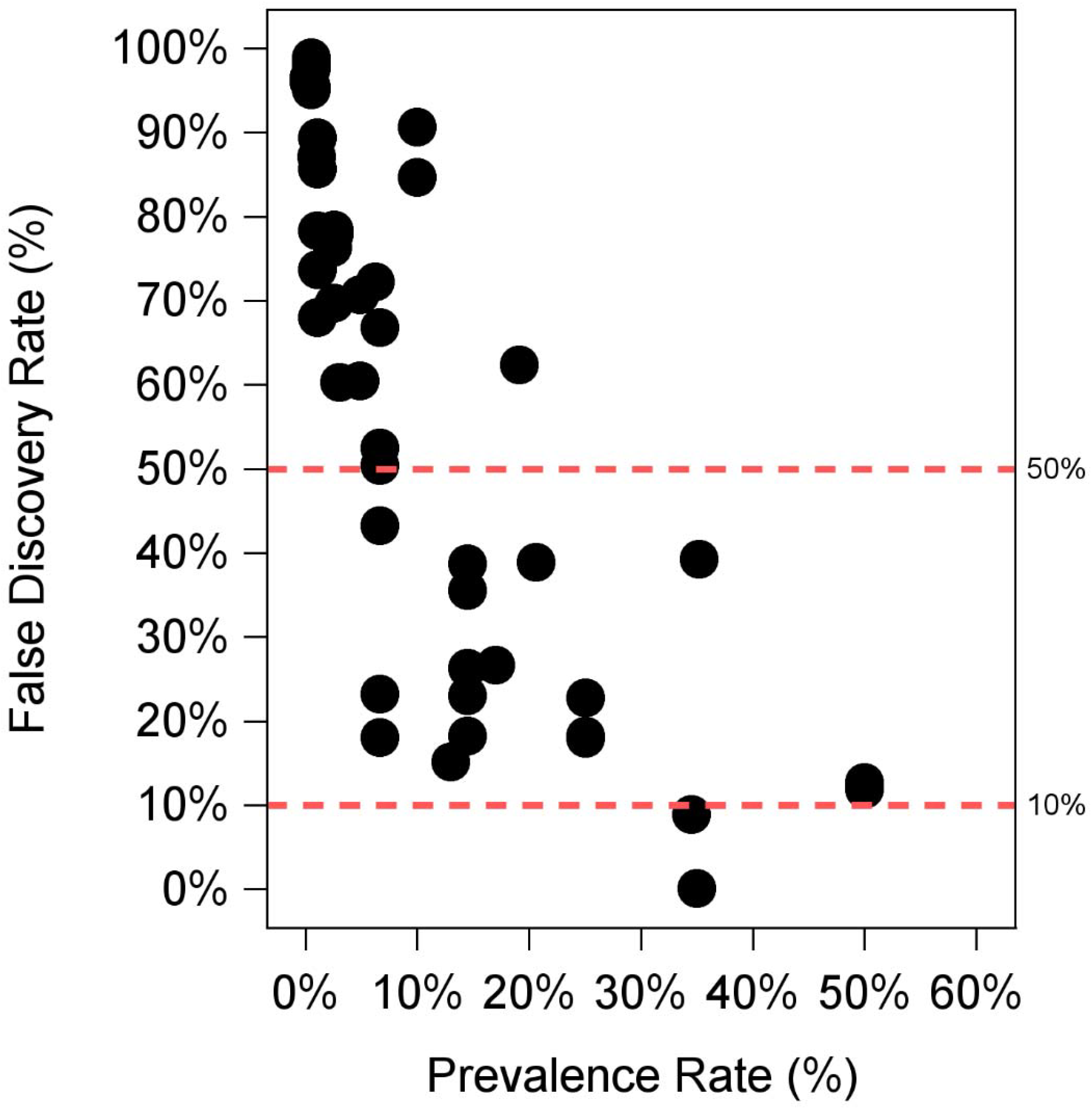
Relationship between False Discovery Rate and Clinical Prevalence. The Y-axis is the False Discovery Rate and the X-axis is the Prevalence Rate. Red dashes note the FDR at 50% and 10% for reference.

For example, in one summary (K243145), even though the sensitivity and specificity of an LVO detector were reported as 90.6% and 88.8%, respectively, the FDR is predicted to be 70.7% (and PPV 29.3%); that is, out of 100 flags, approximately 71 will be false positives. Its FOR is predicted to be 0.54% (and NPV 99.5%); that is, out of 200 non-flags, approximately 1 represents an LVO that was missed. These estimates were derived using LVO prevalence from the recent literature (4.87%)^14^. For comparison, this performance was largely replicated in a recent real-world case study of AI in LVO in clinical settings, where the observed sensitivity was 100%, specificity was 92%, the FDR was 67% (and PPV 33%), and the FOR was 0% (and NPV 100%) with a prevalence of 4.1% ^7^.

Finally, for two summaries (K250248, K241923), the reported PPV and NPV, the former explicitly (22.2% and 99.80% vs. 21.60% and 99.77%, respectively) and the latter implicitly (91.67% and 97.94% vs. 92.0% and 97.85%, respectively), which were very close to the clinically anticipated values, where the prevalence of the former was predicated on our clinically observed prevalence (2.5% vs. 2.52%) and the latter from the literature (34% vs. 35%)^15^.

## Discussion

Even with an overall mean sensitivity, specificity, and ROC-AUC above 90% for summaries reporting these values, as Table 3 demonstrates, the number of false positives (FDR) will likely far exceed what clinical practices and radiologists will anticipate for FDA-authorized devices^7^, save for perhaps a few applications, such as fracture, which has a high prevalence in the literature (50%)^16^. Fortunately, as we have demonstrated here, most public 510(k) summaries reported enough information (i.e., sensitivity and specificity) so that clinical practices can derive the PPV (and FDR) and NPV (and FOR) appropriate for their local practice using these devices.

Though the calculation may be straightforward^17^ (see supplemental materials, Equations 1-4), these resulting PPV (and FDR) and NPV (and FOR) estimates assume 1.) that the vendor-reported sensitivity and specificity estimates generalize to the clinical population in question for a local practice^17,18^, and likewise, 2.) that the estimate of local prevalence, either using their own data or from the literature, is approximately close to the actual local population prevalence.Here, we demonstrate that although most public submission summaries provided sufficient data to accomplish these calculations, these summaries had some shortcomings.

There was little standardization as to what and how information was presented. Although most summaries reported sensitivity and specificity, not all did; few reported only the ROC-AUC (K241747, K242652, K240697) and two proved none of these (K241770, K243688). It is worth noting that one summary (K241770) provided the sensitivity and specificity of radiologists when using the device. Although this may seem sensible, this is, by definition, not the diagnostic performance of the device itself but rather the performance of the radiologist and the device (and its effect on the radiologist), a valid but separate question of diagnostic performance^19^.

Although many summaries provided enough information to calculate the PPV (and FDR) and NPV (and FOR), these calculations likely would have yielded inflated PPV (and deflated FDR) and deflated NPV (and inflated FOR) values due to the use of enriched datasets (unless corrected for real-world prevalence). Nonetheless, if a practice wanted to anticipate the PPV (and FDR) and NPV (and FOR) of a device, they should use these values from the summary only if (1) the summary provides the prevalence of the test dataset and it is similar to that of the practice’s own prevalence or (2) the practice calculates the PPV (and FDR) and NPV (and FOR) using sensitivity, specificity, and their own population’s real-world prevalence (or a close proxy). We note that many summaries did not explicitly report test set prevalence.

In addition, providing a single pair of sensitivity and specificity is also very limiting, namely because it provides diagnostic performance at only a single threshold. This threshold is oftentimes selected using Youden’s J, which mathematically optimizes sensitivity and specificity^20^. However, the mathematically optimal solution for sensitivity and specificity may not always translate into the optimal clinical solution, an issue of the PPV and NPV. That is, the cost of a false positive and the cost of a false negative may need to be weighed differently, along with the frequency of these errors due to prevalence^18,19,21–23^. However, one summary (K241831) reported sensitivity and specificity at “high sensitivity” and “high specificity” thresholds, a good first step in intuiting different threshold for different clinical needs^22^, and another (K250831) provided sensitivity and specificity at multiple thresholds, thus providing practices information to decide the threshold that is best for their practice given their population^17,19,21–23^.

Very few summaries explicitly reported PPVs and NPVs (n=6), and n=4 provided them based on prevalences that likely far exceed clinical prevalence, rendering these estimates very likely inflated, as demonstrated in Table 3 when comparing the vendor reported values with the clinical-anticipated values. Interestingly, two summaries (K251406, K250248) calculated the PPV and NPV using a smaller base rate (both 2.5%, which we back-engineered) relative to the prevalence of their test set (40.3% and 48.7%, respectively), though neither of these base rates were explicitly reported. Even more interesting, even though a base rate of 2.5% still exceeded the base rate from clinical data (0.32%) for aortic dissection (K251406), the other summary for pulmonary embolism (K250248) used a prevalence almost identical to our own estimate (2.52%) and as a result, this summary reported PPV and NPV were practically identical to our clinically anticipated values.

Based on these observations, the following recommendations are made:

1. Sensitivity and specificity should always be reported along with test set prevalence. The reason for this is simple: along with local or published prevalence, these values allow practices to calculate and thus anticipate an AI’s PPV (and FDR) and NPV (and FOR) for their local population. And sensitivity and specificity should always be reported for standalone devices (not with readers) so the error rates of the device itself may be derived.
2. Sensitivity and specificity should be reported for many, if not all, thresholds of AI scores or outputs when possible. Along with local or published prevalence, this permits local practices to choose a threshold based on their risk appetite and tolerance^17,19–23^. Specifically, by being able to quantify the number of false positives (FDR) and false negatives (FOR), a practice can then weigh the cost and benefit of each type of error with the number of each type of error, choosing a threshold that makes sense clinically, ethically, and financially^17^. We note that another way to provide this information is to include the ROC curve itself as a figure, with sensitivity and 1-specificity coordinate pairs.
3. If reporting PPV and NPV, the base rate used to calculate each should be explicitly reported and should reflect a clinically defensible and cited prevalence from the literature. This is not to say that test sets should not be enriched to improve estimates of sensitivity, but rather that the PPV and NPV should be calculated using Bayes’ theorem with these sensitivity and specificity estimates, along with one or more defensible prevalence estimates from the literature. This allows practices to evaluate if the PPVs and NPVs reflect their own clinical practice.
4. Summaries should explicitly report the prevalence of the test set and note if it is artificially high or not. If it is artificially high, then they should include a warning that PPV (and FDR) and NPV (and FOR) should not be calculated using this prevalence.

### Limitations

The present study has limitations. First, this is not an exhaustive review of the prevalence literature for these pathology targets, nor is it meant to be. For example, the prevalence of prostate cancer will vary based on a general screening population or a PSA-selected population, or pleural effusion will be greater in the ICU compared to outside the ICU. Likewise, the clinical prevalence observed here reflects a large, multisite practice and likely does not generalize to all practices and their patient populations. Whether these prevalence rates are debatably too high or too low for a given practice and its patient population only reinforces the central point of the paper: to anticipate the number of false positives and false negatives when using an AI system, practices must use a prevalence rate that reflects their local practice and its local patient population.

Relatedly, the values observed here are not static but moving targets, even for the same patient practice using the same AI system. AI algorithms will change over time (e.g., updates), and local prevalence rates can and likely will also change over time with changes to patient mix, care practices, or other factors. Therefore, performance estimates need to be reviewed and updated periodically to ensure the estimated FDR and FOR reflect the population as best as possible^23^. In addition, as the specificity of AI systems improve, or approaches 100%, with updates, the FDR will decrease, even for small prevalence rates.

## Data Availability

All data are presented in Table 3 and can also be found one the FDA 510k summary website

https://www.accessdata.fda.gov/scripts/cdrh/cfdocs/cfPMN/pmn.cfm

## Supplemental Materials (Equations 1-4)

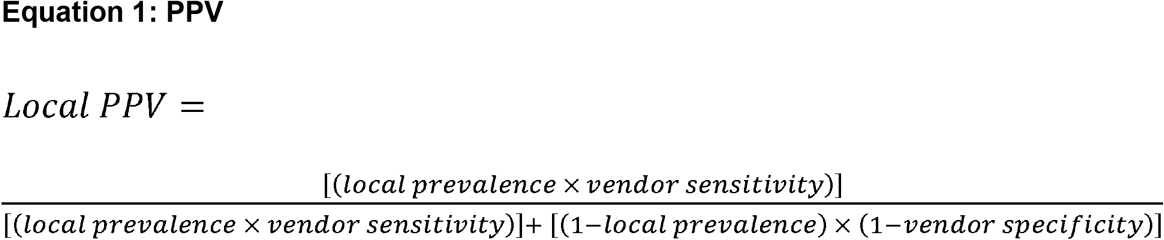

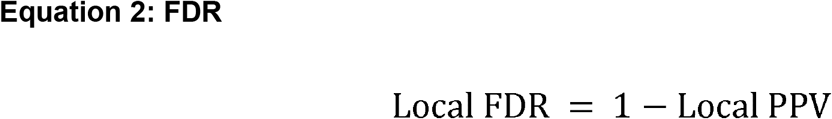

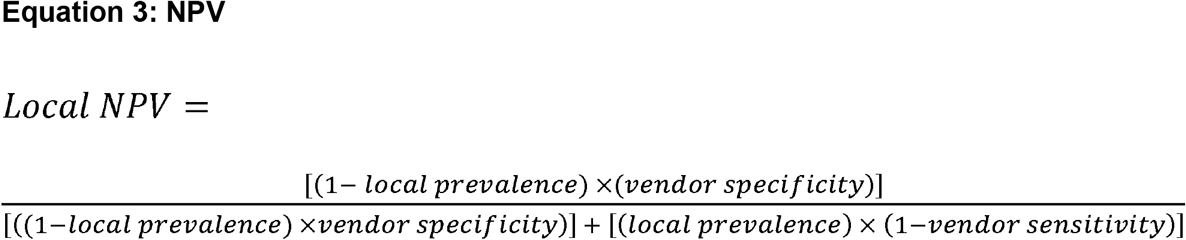

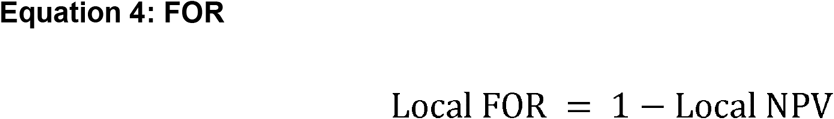

